## Supplement for "Clinical evaluation of the Diagnostic Analyzer for Selective Hybridization (DASH): a point-of-care PCR test for rapid detection of SARS-CoV-2 infection"

*Nasal Swab Stability*

The stability of swabs stored at room temperature and refrigerated and frozen conditions before testing with the Diagnostic Analyzer for Selective Hybridization (DASH) SARS-CoV-2/S were evaluated. A known positive residual patient specimen was used to seed the swabs in this study. A nasopharyngeal specimen in viral transport media (VTM) was estimated to contain $10^5$ copies/μl by comparing the Ct of the diluted specimen to Cts of known concentrations of heat-inactivated virus (USA_WA1/2020, lot 70037779, BEI Resources, NR-52286). The specimen was serially diluted in negative nasal matrix to 15 copies/μl such that 0.01% of the specimen was from the original VTM specimen and the remaining volume consisted of negative nasal matrix. 20 μl volume was applied to each swab to create samples of 3X LOD (300 copies/swab). For negative swabs, 20 μl volume of negative nasal matrix was applied to each swab.

Control swabs (time=0) were tested according to the DASH SARS-CoV-2/S Instructions for Use (IFU) and Quick Reference Guide (QRG). DASH results (positive or negative result and Ct values) were recorded. The swabs were subsequently incubated at room temperature (22°C), 30°C, 4°C, and -80°C for up to 7 days (see Table below). At specified time points the swabs were allowed to equilibrate to room temperature and tested according to the DASH IFU and QRG. Three or four replicates per time point were tested. DASH results (positive or negative and Ct values) were recorded.

Supplemental Table. Testing Conditions for Swab Specimen Stability Studies

| **-80C** | **4C** | **22C** | **30C** |
|---|---|---|---|
| 0 days | 0 days | 0 days | 0 days |

| | | | |
|---|---|---|---|
| 7 days | 1 day | 1 hour | 1 hour |
| | 2 days | 2 hours | 2 hours |
| | 3 days | 4 hours | 4 hours |
| | 4 days | 8 hours | 8 hours |
| | 5 days | 1 day | 1 day |
| | 6 days | 2 days | 2 days |
| | 7 days | 4 days | 4 days |

Supplemental Figure. Days swabs contrived with 300 copies/swab of known positive patient sample were incubated versus percent change of average Ct from time zero.

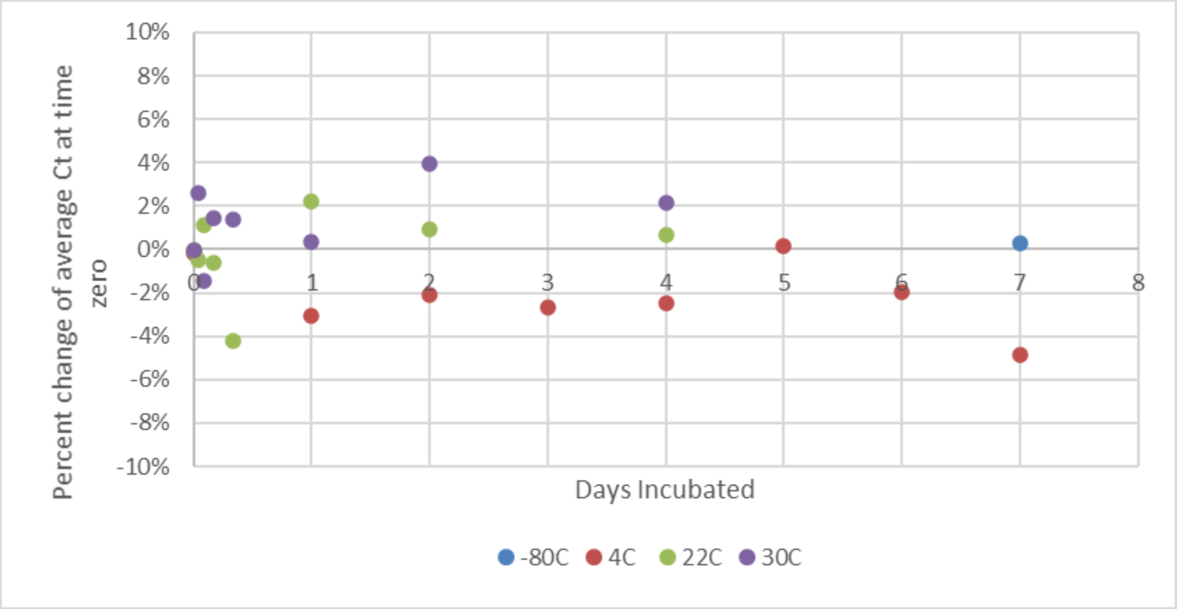


Percent change from the average Ct at time 0 was plotted versus hours of incubation (Supplemental Figure). At each time point, there was less than 10% difference from the Ct value at time 0. This study supports specimen stability for the purposes of the clinical study.
